# More than smell - COVID-19 is associated with severe impairment of smell, taste, and chemesthesis

**DOI:** 10.1101/2020.05.04.20090902

**Authors:** Valentina Parma, Kathrin Ohla, Maria G. Veldhuizen, Masha Y Niv, Christine E Kelly, Alyssa J. Bakke, Keiland W. Cooper, Cédric Bouysset, Nicola Pirastu, Michele Dibattista, Rishemjit Kaur, Marco Tullio Liuzza, Marta Y. Pepino, Veronika Schöpf, Veronica Pereda-Loth, Shannon B Olsson, Richard C Gerkin, Paloma Rohlfs Domínguez, Javier Albayay, Michael C. Farruggia, Surabhi Bhutani, Alexander W. Fjaeldstad, Ritesh Kumar, Anna Menini, Moustafa Bensafi, Mari Sandell, Iordanis Konstantinidis, Antonella Di Pizio, Federica Genovese, Lina Öztürk, Thierry Thomas-Danguin, Johannes Frasnelli, Sanne Boesveldt, Özlem Saatci, Luis R. Saraiva, Cailu Lin, Jérôme Golebiowski, Liang-Dar Hwang, Mehmet Hakan Ozdener, Maria Dolors Guàrdia, Christophe Laudamiel, Marina Ritchie, Jan Havlícek, Denis Pierron, Eugeni Roura, Marta Navarro, Alissa A. Nolden, Juyun Lim, KL Whitcroft, Lauren R Colquitt, Camille Ferdenzi, Evelyn V Brindha, Aytug Altundag, Alberto Macchi, Alexia Nunez-Parra, Zara M. Patel, Sébastien Fiorucci, Carl M Philpott, Barry C. Smith, Johan N. Lundström, Carla Mucignat, Jane K. Parker, Mirjam van den Brink, Michael Schmuker, Florian Ph.S Fischmeister, Thomas Heinbockel, Vonnie D.C. Shields, Farhoud Faraji, Enrique Santamaría, William E.A. Fredborg, Gabriella Morini, Jonas K. Olofsson, Maryam Jalessi, Noam Karni, Anna D’Errico, Rafieh Alizadeh, Robert Pellegrino, Pablo Meyer, Caroline Huart, Ben Chen, Graciela M. Soler, Mohammed K. Alwashahi, Antje Welge-Lüssen, Jessica Freiherr, Jasper H. B. de Groot, Hadar Klein, Masako Okamoto, Preet Bano Singh, Julien W. Hsieh, GCCR Group Author, Danielle R Reed, Thomas Hummel, Steven D. Munger, John E. Hayes, Olagunju Abdulrahman, Pamela Dalton, Carol H. Yan, Vera V. Voznessenskaya, Jingguo Chen, Elizabeth A. Sell, Julie Walsh-Messinger, Nicholas S. Archer, Sachiko Koyama, Vincent Deary, S. Craig Roberts, Hüseyin Yanik, Samet Albayrak, Lenka Martinec Nováková, Ilja Croijmans, Patricia Portillo Mazal, Shima T. Moein, Eitan Margulis, Coralie Mignot, Sajidxa Mariño, Dejan Georgiev, Pavan K. Kaushik, Bettina Malnic, Hong Wang, Shima Seyed-Allaei, Nur Yoluk, Sara Razzaghi-Asl, Jeb M. Justice, Diego Restrepo

## Abstract

Recent anecdotal and scientific reports have provided evidence of a link between COVID-19 and chemosensory impairments such as anosmia. However, these reports have downplayed or failed to distinguish potential effects on taste, ignored chemesthesis, generally lacked quantitative measurements, were mostly restricted to data from single countries. Here, we report the development, implementation and initial results of a multi-lingual, international questionnaire to assess self-reported quantity and quality of perception in three distinct chemosensory modalities (smell, taste, and chemesthesis) before and during COVID-19. In the first 11 days after questionnaire launch, 4039 participants (2913 women, 1118 men, 8 other, ages 19-79) reported a COVID-19 diagnosis either via laboratory tests or clinical assessment. Importantly, smell, taste and chemesthetic function were each significantly reduced compared to their status before the disease. Difference scores (maximum possible change ±100) revealed a mean reduction of smell (−79.7 ± 28.7, mean ± SD), taste (−69.0 ± 32.6), and chemesthetic (−37.3 ± 36.2) function during COVID-19. Qualitative changes in olfactory ability (parosmia and phantosmia) were relatively rare and correlated with smell loss. Importantly, perceived nasal obstruction did not account for smell loss. Furthermore, chemosensory impairments were similar between participants in the laboratory test and clinical assessment groups. These results show that COVID-19-associated chemosensory impairment is not limited to smell, but also affects taste and chemesthesis. The multimodal impact of COVID-19 and lack of perceived nasal obstruction suggest that SARS-CoV-2 infection may disrupt sensory-neural mechanisms.

## Introduction

In late 2019, a new virus, SARS-CoV-2 (Severe Acute Respiratory Syndrome coronavirus strain 2), was reported in Wuhan, China (Zhu et al., 2020). The resulting COVID-19 disease has become a global pandemic with 3.18 million reported cases as of May 1, 2020 (World Health Organization, 2020). When assessing SARS-CoV-2 infection, clinicians initially focused on symptoms such as fever, body aches, and dry cough. However, emerging reports suggest sudden olfactory loss (anosmia or hyposmia) may be prevalent in patients with COVID-19 (Menni et al., 2020; Vetter et al., 2020). Olfactory disorders have long been associated with viral upper respiratory tract infections (URI) that cause the common cold and flu, including influenza and parainfluenza viruses, rhinoviruses, and other endemic coronaviruses (Soler et al., 2020). Taste disorders have been known to occur during and after respiratory viral infection, as well (Hummel et al., 2011). One case report found anosmia presenting with SARS (Hwang, 2006). Olfactory dysfunction due to viral infections may account for 11-45% of all olfactory disorders excluding presbyosmia (Nordin and Brömerson, 2008). The estimated prevalence of COVID-19-associated olfactory impairment may be higher than in COVID-19-independent postviral olfactory loss; estimations range from 5% to 85% in self-report studies, with differences noted between mild and severe cases (Bagheri et al., 2020; Gane et al., 2020; Giacomelli et al., 2020; Haldrup et al., 2020; Hopkins et al., 2020; Lechien et al., 2020a; 2020b; Mao et al., 2020; Menni et al., 2020; Yan et al., 2020a; 2020b). When psychophysical odor identification tests are used, this prevalence ranges from 76% in Europe using the Sniffin’ Sticks (Lechien et al., 2020b) to 98% in Iran using the UPSIT (Moein et al., 2020), though the severity of COVID-19 in these study cohorts may not be representative of the larger population. These anecdotes, pre-prints, letters, and peer-reviewed reports (for a review see, Pellegrino et al., in press), describe chemosensory disturbances in COVID-19 with characteristics that are similar to those seen in common URIs, such as isolated sudden onset of anosmia (Gane et al., 2020), occurrence of anosmia in mild or asymptomatic cases of COVID-19 (Hopkins et al., 2020), and loss of taste (Lechien et al., 2020a; Yan et al., 2020a). As of May 13, 2020, the European Centre for Disease Prevention and Control, the World Health Organization and the following countries or regions have listed smell loss as a symptom of COVID-19: Argentina, Chile, Denmark, Finland, France, Italy, Luxembourg, New Zealand, Singapore, South Africa, Slovenia, Spain, Switzerland, The Netherlands, and the United States of America (U.S.A.); many other countries or regions have not yet officially acknowledged smell loss as a symptom of COVID-19. To date, quantitative studies to determine the extent and detail of broad chemosensory changes in COVID-19 are rare, with the exception of two recent studies: Iravani et al. (2020), assessed odor intensity in a group of Swedish respondents, while Moein et al. (2020) tested a small sample of hospitalized Iranian patients with the University of Pennsylvania Smell Identification Test. We use three separate sensory modalities – smell, taste and chemesthesis – to sense our chemical environment in daily life. The olfactory system (smell) detects volatile chemicals through olfactory sensory neurons in the nasal cavity. Odors in the external environment are sampled through the nostrils (orthonasal olfaction), while odors coming from food or drink in the mouth are sampled via the nasopharynx (retronasal olfaction). The gustatory system (taste) responds to non-volatile compounds in the mouth that elicit sensations of sweet, salty, bitter, sour and umami (savory). Finally, chemesthesis detects other chemicals, often found in herbs or spices, that evoke sensations like burning, cooling or tingling.

While taste has occasionally been explored with respect to COVID-19 (e.g., Giacomelli et al., 2020; Yan et al., 2020), chemesthesis remains unexamined in recent studies, despite anecdotal reports that it may be similarly compromised in persons with COVID-19. Smell, taste, and chemesthesis are often conflated, mostly because they produce a single experience of flavor during eating (Rozin, 1982; Spence et al., 2014; Duffy and Hayes, 2019; Hayes, 2019), and patients often report a loss of taste when in fact they are experiencing a loss of retronasal olfaction. Nevertheless, the olfactory and gustatory systems, along with parts of the somatosensory system that conveys chemesthesis, are separate sensory systems with distinct peripheral and central neural mechanisms (Shepherd, 2006; Green, 2012). To date, the impact of COVID-19 on each of these three chemosensory modalities remains poorly understood.

Chemosensory disturbances can result in quantitative reductions in smell or taste (i.e., anosmia/hyposmia and ageusia/hypogeusia, respectively), or as qualitative changes (e.g., distortions of smell and taste, termed parosmia and dysgeusia, or phantom sensations, termed phantosmia and phantogeusia). These key distinctions have been neglected in previous reports. Because these phenomena are not necessarily correlated and have different mechanisms (Holbrook et al., 2005; Iannilli et al., 2019; Reden et al., 2007), understanding how COVID-19 impacts chemosensation in both quantitative and qualitative ways should provide important insights into the mechanisms by which the SARS-CoV-2 virus affects the chemical senses.

Ideally, validated testing of chemosensory function would be combined with a review of a patient’s medical records, including laboratory test results (from viral swab or serology, “Lab Test”) to confirm the infectious agent. Due to limited laboratory test availability in many countries, the necessity in some medical settings for social distancing, and a potentially large number of asymptomatic or mild cases, it has been impractical or impossible to conduct such chemosensory testing for many individuals with COVID-19. Additionally, in many countries where testing resources are limited, laboratory testing has been limited to the most severe cases. Another diagnosis method is a clinical assessment by a medical professional (“Clinical Assessment”), either in-office or remotely via tele-medicine. Thus, the method of diagnosis – Lab Test versus Clinical Assessment – may be associated with differences in symptom severity, including severity of chemosensory impairments. To account for possible differences in the severity of infection as well as the availability of diagnosis options across countries, we collected information on diagnosis methods and compared chemosensory function between participants diagnosed with Lab Test vs. Clinical Assessment.

Given all the issues raised above, we deployed a crowd-sourced, multilingual, online study with a global reach (as of May 1, 2020 deployed in 27 languages); this survey has the potential to provide reproducible data from a large number of participants around the world. In this pre-registered report, we present data from 4039 participants who reported a COVID-19 diagnosis either via Lab Test or Clinical Assessment and who completed the questionnaire during the first 11 days the study was available online. Here we address two main research questions. First, we asked what chemosensory changes are observed in participants with COVID-19, compared to before illness (i.e., within participants). Next, we asked whether the two diagnostic groups differ in chemosensory changes (i.e., between participants). For both diagnosis methods, we observed significant quantitative changes in smell, taste, and chemesthesis with COVID-19. Most chemosensory loss could not be accounted for by self-reported nasal obstruction, a factor commonly associated with diminished smell in other upper respiratory diseases (Doty, 2001). Further, we found little incidence of qualitative changes in olfactory function, with only a small percentage of participants reporting distorted smells (consistent with parosmia) or phantom smells (consistent with phantosmia). Together, these results provide an initial assessment of comprehensive chemosensory impairments associated with COVID-19.

## Method

### Preregistration

We preregistered our hypotheses and analyses on April 19, 2020, at 12:20 AM Eastern Daylight Time (EDT), before the data became available (data reflected questionnaires submitted between April 7, 2020 6:00AM EDT and April 18, 2020 at 8:34 AM EDT) (Veldhuizen et al., 2020). In line with the pre-registration, and according to the Sequential Bayes Factor Design *(section 2.3)*, one of the authors (AJB) not involved in the development of the pre-registration queried the database to check whether the minimum number of participants per group was reached. The data reported in this manuscript, along with analysis scripts, will be available at OSF (https://osf.io/a3vkw/) upon the acceptance of the manuscript. The project is structured according to the research compendium created with the *rrtools* package (Marwick, 2019). The presented analyses are as pre-registered, unless specified otherwise.

### The GCCR core questionnaire

The GCCR questionnaire [Supplementary Materials, and included in the list of research tools to assess COVID-19 by the NIH Office of Behavioral and Social Sciences Research (OBSSR, Anonymous, 2020)], measures self-reported smell, taste, and chemesthesis function as well as nasal blockage in participants with respiratory illness, including COVID-19, within the two weeks prior to completing the questionnaire. It was created iteratively through a crowdsourced approach with a preliminary period of development and commentary across an international group of chemosensory experts, clinicians and patients advocates. Relevant to the scope of the present manuscript, participants were asked to quantify their ability to smell, taste, and perceive cooling, tingling and burning sensations (chemesthesis) before and during the COVID-19, on separate, horizontally-presented, 100-point visual analogue scales (VAS). Participants were also asked to quantify their perceived nasal obstruction on a 100-point VAS with “not at all blocked” and “completely blocked” as anchors. Framing the questions in terms of ability, rather than intensity, was driven by the desire to be readily understood by participants without additional training or instructions and was informed by spontaneous patient reports, internet search trends and in dialogue with patient advocates (e.g., we implicitly separated taste / chemesthesis experienced in the mouth from orthonasal smell as experienced in the nose, in full alignment with the ecological framework first proposed by Gibson in 1966. Specifically, for taste, we stated “The following questions are related to your sense of taste. For example, sweetness, sourness, saltiness, bitterness experienced in the mouth.” For chemesthesis, we stated “The following questions are related to other sensations in your mouth, like burning, cooling, or tingling. For example, chili peppers, mint gum or candy, or carbonation. In both cases, we were orienting participants toward sensations that are experienced in the mouth. In contrast, for smell, we stated “These questions relate to your sense of smell (for example, sniffing flowers or soap, or smelling garbage) but not the flavor of food in your mouth.” The within-subject nature of the present design precludes a need for more sophisticated scaling methods than VAS (Kalva et al., 2014). Although participants were not randomly assigned to the two diagnostic groups, the groups may be considered *as if* random when it comes to adjective interpretation / scale usage, thereby within Bartoshuk’s arguments for using a VAS across group comparisons (Bartoshuk et al., 2003). Please refer to Figure S2 for a list of the questions analyzed in the present work.

Participants were also asked to report demographic information (i.e., year of birth, gender, and country of residence) as well as information related to their COVID-19 diagnosis and their respiratory illness-related symptoms, including smell and taste, in check-all-that-apply (CATA) format. We summarized the questions used in the present study in Supplementary Figure S1. Please refer to the full questionnaire, included in the Supplementary materials, for question order and the labels on the anchors of each question.

The questionnaire was implemented in 10 languages as of April 18th, 2020 (the date on which the database was last queried for this report): English, French, German, Italian, Japanese, Kannada, Norwegian, Spanish, Swedish, and Turkish. Our translation protocol was modeled after the process developed by the Psychological Science Accelerator (Moshontz et al., 2018). Briefly, translations of the original English questionnaire involved three steps: i) the original (English) questionnaire was translated to the target language by independent translators, resulting in Translation Version A; ii) Version A was translated back from the target language to English by a separate group of independent translators, resulting in Version B; iii) Versions A and B were discussed among all translators, with the goal of resolving potential discrepancies between the two versions, resulting in the final Version C. All questionnaires in all languages were then implemented in Compusense Cloud, Academic Consortium (Guelph, Ontario), a secure cloud-based data collection platform with multilingual support. Please refer to the supplementary materials for the full survey (Supplementary Materials) and to the questions from the survey analyzed in the present work (Figure S1).

### Study design

This study compares self-reported quantitative changes (during vs. before the illness) in smell, taste, chemesthesis, and nasal obstruction as well as qualitative changes in smell and taste between two groups of respondents: those who reported a COVID-19 diagnosis as a result of an objective test such as a swab test (“Lab Test”) or those who reported a diagnosis from clinical observations by a medical professional (“Clinical Assessment”). Given the lack of effect size estimates in the literature, we employed a Sequential Bayes Factor Design (SBFD) that allows optional stopping with unlimited multiple testing (Schönbrodt et al., 2017). Specifically, we used a SBFD with a minimal number of participants and a temporal stopping rule to increase the probability of obtaining the desired level of evidence and to reduce the probability of obtaining misleading evidence. The desired grade of relative evidence for the alternative vs. the null (BF_10_) hypothesis is set at BF_10_ > 10 (strong evidence) for H_1_ and BF_01_ > 6 (moderate evidence) for H_0_. We derived the minimal N_min_ = 480 per group to start SBFD through a Bayes Factor Design Analysis (BFDA) for fixed-n designs (Schönbrodt and Wagenmakers, 2018) for a two-independent-sample, two-sided testing, and a conservative Cohen’s D = 0.2 with 80% power of reaching a BF_10_ > 10 and a BF_01_ > 6 with a default prior. Our stopping rule follows a temporal criterion (data collection until April 18, 2020, 8:34 AM EDT) and N_min_. BF computation continues with every 20 participants added in the slowest accumulating group at a time until the thresholds of H_1_ or H_0_ are reached.

### Study setting

Participation in this online study was voluntary and participants received no remuneration. Inclusion criteria were: consent to participate, age 19 years and older (based on birth year), and any form or suspicion of respiratory illness in the past two weeks. Participants were asked about their year of birth and the onset of their illness during the survey to confirm the inclusion criteria, and the survey terminated for non-eligible participants via branching logic. The nature of the questionnaire necessitated at least some secondary education in terms of language and distribution method (web survey) as well as internet access. The protocol complies with the revised Declaration of Helsinki and was approved as an exempt study by the Office of Research Protections at The Pennsylvania Study University (Penn State) in the U.S.A. (STUDY00014904). The questionnaire was distributed globally in the different languages through traditional (i.e., print, television, radio) and social media (e.g., Twitter, Facebook), the website of the Global Consortium for Chemosensory Research (GCCR; https://gcchemosensr.org), flyers, professional networks, and word of mouth. All data were collected from a non-representative, convenience sample via Compusense Cloud, which is compatible with use on a smartphone, tablet, laptop, or desktop computer. Data collection was compliant with privacy laws in the U.S.A. and the European Union [including California and General Data Protection regulation (GDPR) rules].

### Participants

At the close of data collection on April 18, 2020, 4039 participants with a diagnosis of COVID-19 completed the ratings for smell, taste, chemesthesis ability, and nasal obstruction before and during their recent illness and were included in the present study. Participants who did not complete all ratings as mentioned above and/or gave inconsistent responses in three questions that addressed changes in smell perception (specifically, selecting changes in smell in “Have you had any of the following symptoms with your recent respiratory illness or diagnosis?”, reporting a difference of at least 5 points in “Rate your ability to smell before your recent respiratory illness or diagnosis” and/or select at least one answer at the question “Have you experienced any of the following changes in smell with your recent respiratory illness diagnosis?”) or reported an age above 100 (n = 1) were excluded from the sample. Of those included in the final sample, 2913 were women, 1118 were men, 3 were other and 5 preferred not to say. Overall the age of the participants ranged from 19 to 79 years old (mean ± sd: 41.38 ± 12.20 years old).

Here, we will compare respondents from two diagnostic groups: (a) participants who reported that their COVID-19 diagnosis was confirmed via objective Lab Test (N = 1402: 1064 F, 335 M; age mean ± sd: 40.73 ± 12.29 years old) compared with (b) participants who reported that their COVID-19 diagnosis was obtained via clinical observation by a medical professional (N = 2637: 1849 F, 783 M; age mean ± sd: 41.72 ± 12.14 years old). Based on self-report, respondents indicated they resided in the following countries: Algeria, Argentina, Australia, Austria, Belgium, Brazil, Canada, Colombia, Costa Rica, Czech Republic, Denmark, Ecuador, Egypt, France, Germany, Greece, Iran, Ireland, Italy, Luxembourg, Morocco, Mexico, Netherlands, New Zealand, Norway, Paraguay, Portugal, Romania, Russia, Singapore, Slovenia, South Africa, Spain, Sweden, Switzerland, Thailand, Tunisia, Turkey, UK, United Arab Emirates, U.S.A. Figure 1 illustrates the derivation of the sample presented here.

**Figure 1.**
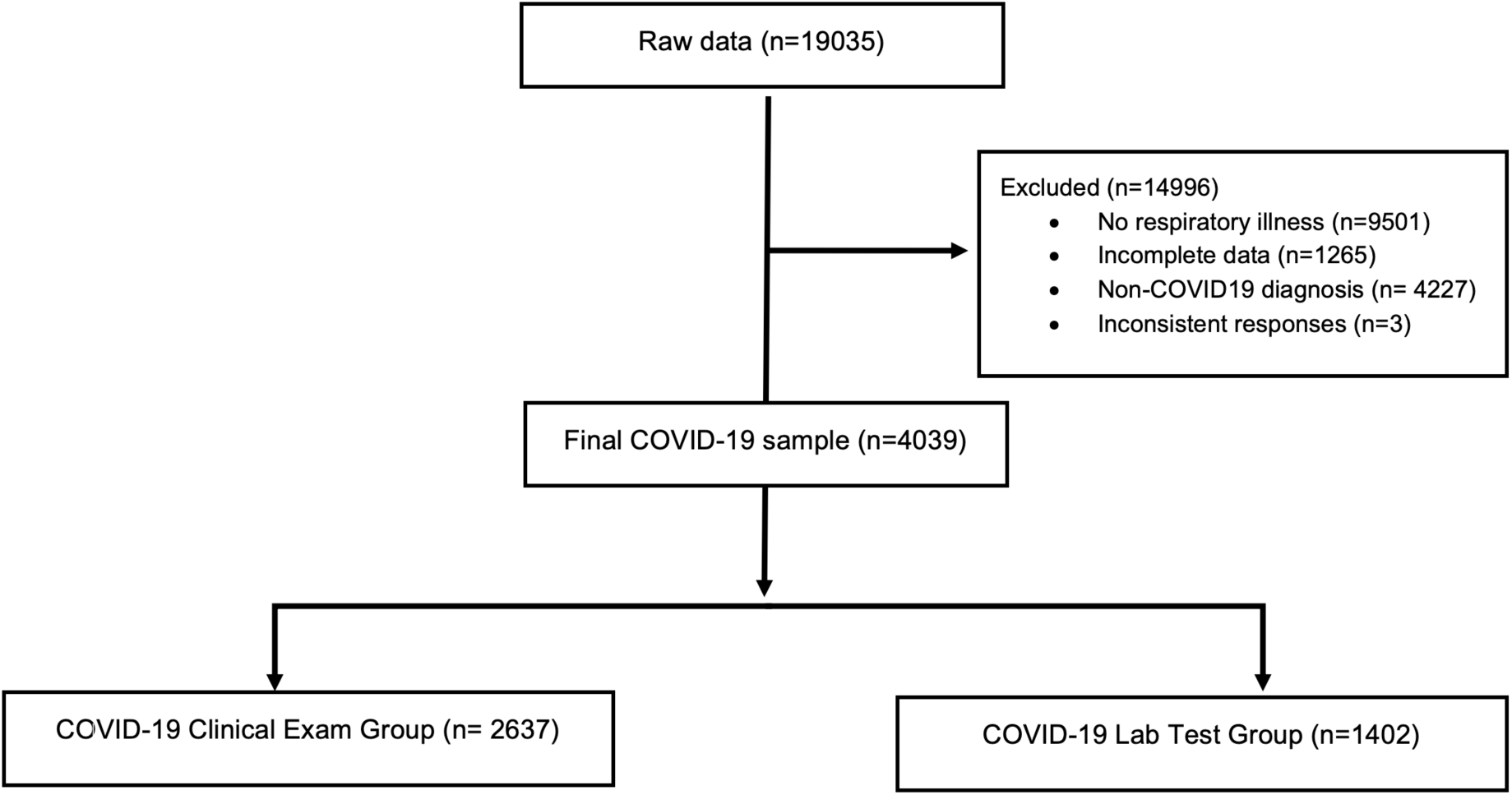
Flow diagram showing the selection of individual observations included in the reported analysis as well as the observations that were excluded.

### Statistical analysis

All analyses were performed in R (Team R Core Development, 2013) via RStudio. The scripts along with information on the computational environment and dependencies will be found, upon acceptance of the manuscript, at https://osf.io/a3vkw/. Information on the computational environment and dependencies used is also shared for future reproducibility. The code will be also available on GitHub at https://github.com/GCCR/GCCR001, and will include a Jupyter notebook replicating the core analyses in Python.

To test our hypotheses (H_0_: no difference between groups; H_1_: difference between groups) in this between-participant SBFD, we conducted a Bayesian linear regression with the *ImBF* function from the *BayesFactor* package (Morey and Rouder, 2018) to detect changes (during minus before COVID-19) in smell, taste and chemesthetic abilities as well as nasal obstruction. Data report the Bayes factor and no proportional error estimate on the Bayes factor since they were all lower than 2.07e-05. We used the default Cauchy prior on the effect sizes under the H_1_ as the scale parameter spread which was set at its default value of r = sqrt(2)/2. We performed robustness (sensitivity) checks by adjusting the Cauchy distribution to *r* = 0.5 and *r* = 1 to assess how the choice of prior affects the conclusions drawn from the analysis. We first assessed whether the model provides evidence in favor of H_1_ or H_0_. To interpret the strength and the direction of those effects, we sampled from the models’ posterior distributions (iterations = 1e4). Please refer to the pre-registration and the analysis script (see above) for further details. As reported in Table 1, the interpretation of the Bayes factors BF_10_ follows the classification scheme proposed by Lee and Wagenmakers (2013) and adjusted from (Jeffreys, 1961).

**Table 1.** *Interpretation of the Bayes factors BF_10_ follows the classification scheme proposed by Lee and Wagenmakers (2013) and adjusted from Jeffreys (1961)*.

| Bayes Factor | Evidence Category |
| --- | --- |
| >100 | Extreme evidence for $H_1$ |
| 30 -100 | Very strong evidence for $H_1$ |
| 10 -30 | Strong evidence for $H_1$ |
| 3 - 10 | Moderate evidence for $H_1$ |
| 1 - 3 | Anecdotal evidence for $H_1$ |
| 1 | No evidence |
| 1/3 - 1 | Anecdotal evidence for $H_0$ |
| 1/10 - 1/3 | Moderate evidence for $H_0$ |
| 1/30 - 1/10 | Strong evidence for $H_0$ |
| 1/100 - 1/30 | Very strong evidence for $H_0$ |
| < 1/100 | Extreme evidence for $H_0$ |

### Exploratory non-preregistered analyses

To quantify the association between the reports of (a) parosmia and phantosmia, (b) smell, (c) taste, (d) chemesthesis, and (e) a change in perceived nasal obstruction, we computed a correlation matrix that is visualized with *ggstatsplot* (Patil and Powell, 2018). To assess whether the proportion of parosmia and phantosmia reports differs between groups, we used a two-sample test for equality of proportions with a continuity correction. To characterize the relationship between perceived nasal blockage and chemosensory change, we used a principal component analysis (PCA) using *prcomp* from the R default *stats* package and we plotted the results with functions from the *FactoMineR* package (Lê et al., 2008). Additionally, to test whether different chemosensory function profiles exist in our sample, we performed a cluster analysis. The best clustering scheme was with 3 clusters as determined with *NbCluster* (Charrad et al., 2014), which tests 30 methods that vary the combinations of number of clusters and distance measures for the k-means clustering. Cluster stability was estimated through a bootstrapping approach (100 iterations) with the *bootcluster* package (Yu, 2017).

## Results

### Degree of smell loss during COVID-19

Overall, participants reported a large reduction in the sense of smell (−79.7 ± 28.7 points on the 100-point scale; mean ± sd). Such decrease in the ability to smell was confirmed with extreme evidence (smell change against zero: BF_10_ = 4366.29 ± 0%) and that was similar for both groups (BF_10_ = 2.17 ± 0% inconclusive evidence for a group difference, i.e. H_1_; Figure 2A). The Clinical Assessment group exhibited a larger variance in the ability to smell during the illness as compared to the Lab Test group (Levene test, F_(1,4037)_ = 6.81, p = 0.009; see also the box plots in Figure 2A).

**Figure 2.**
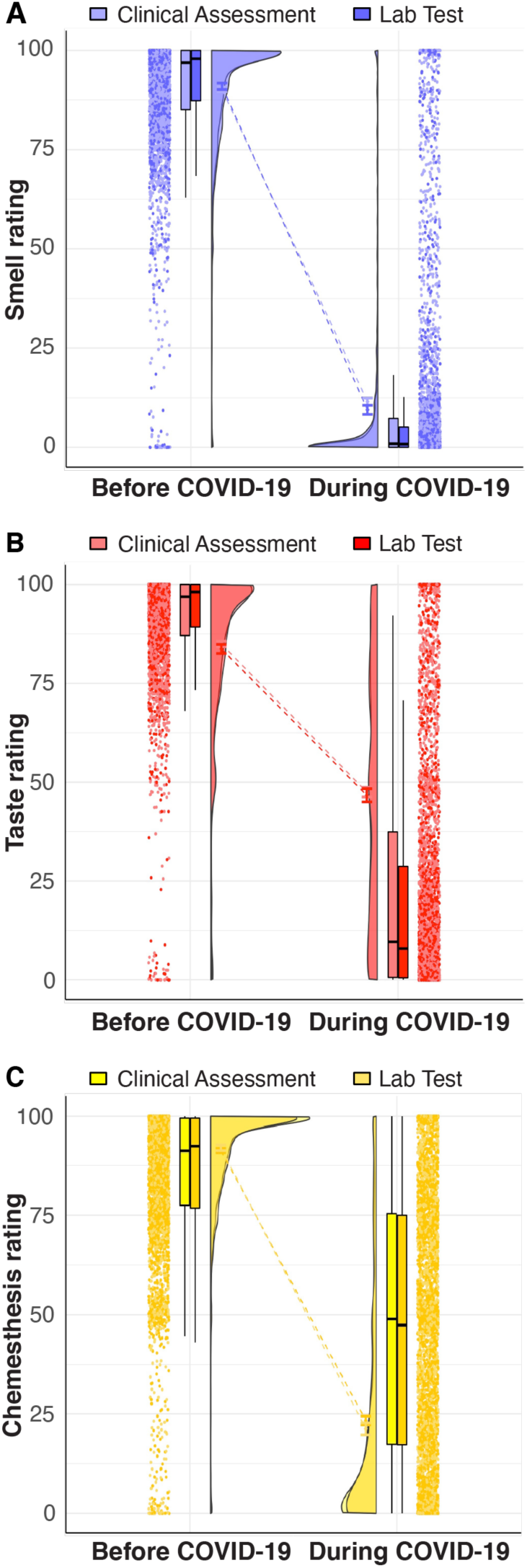
Raincloud plots representing ratings for smell (A), taste (B), and chemesthesis (C) before (left) and during (right) COVID-19. Within each subplot (from left to right), ratings from single participants are displayed as dots. Boxplots show the 1st to 3rd quartiles, the horizontal line denotes the median, and whiskers denote 1.5 times the interquartile range. The density distribution of the data shows the proportions of given ratings. COVID-19 diagnosis is coded such that Clinical Assessment is a lighter shade and Lab Test is a darker shade.

**Table 2.** *Mean and standard deviation (SD) for the ratings of smell, taste, chemesthesis, and nasal obstruction before and during COVID-19 in the Clinical Assessment and Lab Test groups*.

| Variable | Clinical Assessment |  |  |  | Lab Test |  |  |  |
| --- | --- | --- | --- | --- | --- | --- | --- | --- |
|  | Before COVID-19 |  | During COVID-19 |  | Before COVID-19 |  | During COVID-19 |  |
|  | Mean | SD | Mean | SD | Mean | SD | Mean | SD |
| Smell | 90.18 | 14.92 | 11.49 | 24.24 | 90.96 | 15.71 | 9.46 | 22.33 |
| Taste | 91.33 | 13.25 | 23.34 | 29.36 | 92.00 | 14.34 | 21.23 | 28.71 |
| Chemesthesis | 84.96 | 18.74 | 47.48 | 32.17 | 83.72 | 22.1 | 46.68 | 32.2 |
| Nasal Obstruction | 9.83 | 18.41 | 31.67 | 32.11 | 9.35 | 17.89 | 32.67 | 31.62 |

### Smell qualitative changes

Parosmia did not differ significantly between groups 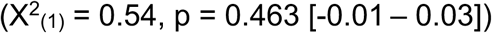 and was reported by 7.77% (205 out of 2637) of participants in the Clinical Assessment and in 7.13% (100 out of 1402) the Lab Test group. Reports of phantosmia, however, did significantly differ between groups 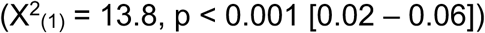: it was reported by 9.44% (249 out of 2637) of participants in the Clinical Assessment and in 6.28% (88 out of 1402) the Lab Test group. Reports of either parosmia or phantosmia negatively correlated with a report of a reduced ability to smell (on VAS) or a total smell loss (reported via CATA). Parosmia and phantosmia positively correlated with changes in smell, taste, and chemesthesis ratings but not with changes in perceived nasal obstruction (Figure 3).

**Figure 3.**
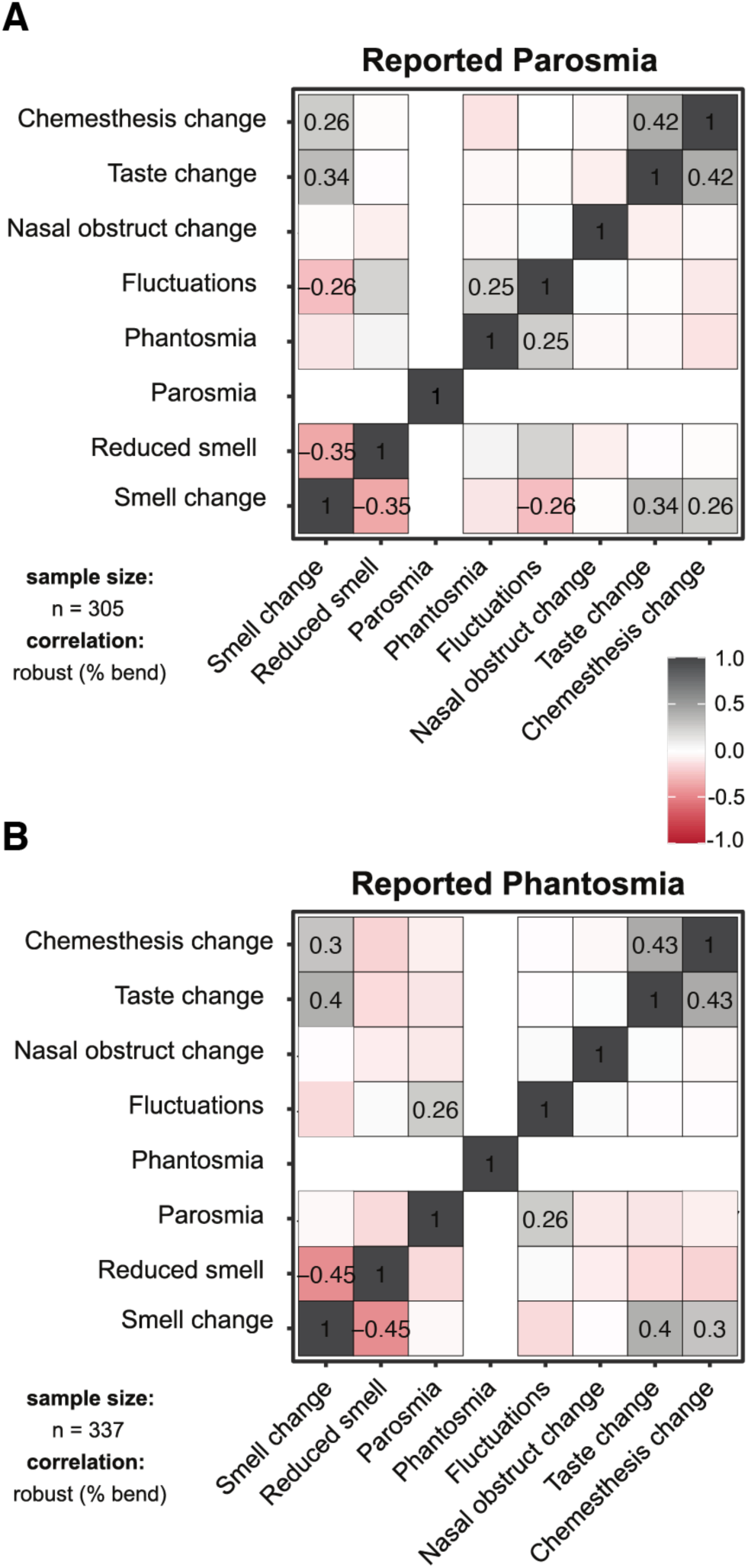
Correlation matrices for individuals who reported parosmia (left, n = 296) and phantosmia (right, n = 324) across groups. The numbers refer to significant correlations at p < 0.001 (Adjustment: Holm).

### Degree of taste loss in COVID-19

Similar to what was seen with smell loss, we observed an overall reduced ability to taste (−69.0 ± 32.6 points; mean ± sd) that was confirmed with extreme evidence (taste change against zero: BF_10_ = 3424.52 ± 0%) and that was similar for both groups (BF_10_ = 0.72 ± 0% suggesting inconclusive evidence for a group difference). The Clinical Assessment group exhibited a larger variance in the ability to taste during COVID-19 as compared to the Lab Test group (Levene test: F_(1,4037)_ = 3.91, p = 0.048; see also the box plots in Figure 2B).

### Taste quality-specific changes

Participants were given the option to report changes in specific taste qualities (i.e., salty, sour, sweet, bitter or umami/savory) as a CATA question. Of all participants, 40% in both groups did not respond, 11% in both groups reported impairment of a single taste quality, and 48% reported impairment of two or more taste qualities (48% in the Clinical Assessment group, 49% in the Lab Test group). Between groups, only umami (savory) taste change was less frequently reported (25%) in the Clinical Assessment group than in the Lab Test group (29%; X2(1) = 7.22, p < 0.007 [-0.07 – −0.01]). No significant differences in the frequency of reporting changes for sweet, salty, bitter or sour taste was evident between groups (Table 3).

**Table 3.**
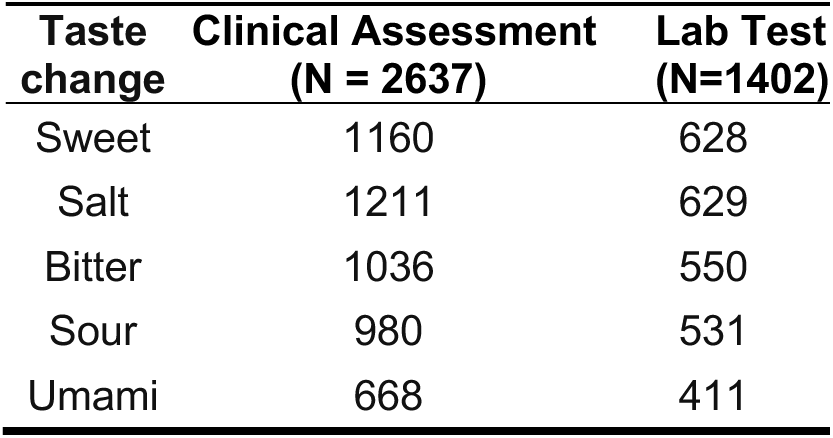
*Frequency of responses, by group, for changes of specific taste qualities during COVID-19*.

| **Taste change** | **Clinical Assessment**<br>**(N = 2637)** | **Lab Test**<br>**(N=1402)** |
| --- | --- | --- |
| Sweet | 1160 | 628 |
| Salt | 1211 | 629 |
| Bitter | 1036 | 550 |
| Sour | 980 | 531 |
| Umami | 668 | 411 |

### Degree of chemesthesis loss in COVID-19

Similar to taste and smell, we observed an overall loss of chemesthetic ability (−37.3 ± 36.2; mean ± sd) that was confirmed with extreme evidence (chemesthetic change against zero: BF_10_ = 1459.98 ± 0%) and that was similar for both groups (BF_01_ = 35.42 ± 0% suggesting strong evidence against a group difference, Figure 2C). The distribution of chemesthetic ability showed a large 95%-CI [-2.82 – 1.88].

### Perceived nasal obstruction in COVID-19

We observed a disease-related change in perceived nasal obstruction that was supported by extreme evidence (nasal obstruction change against zero: BF_10_ = 783.25 ± 0%). No difference in the change in perceived nasal obstruction was found between groups as corroborated by moderate evidence against a group difference (BF_01_ = 14.52 ± 0%; Figure 4A).

**Figure 4.**
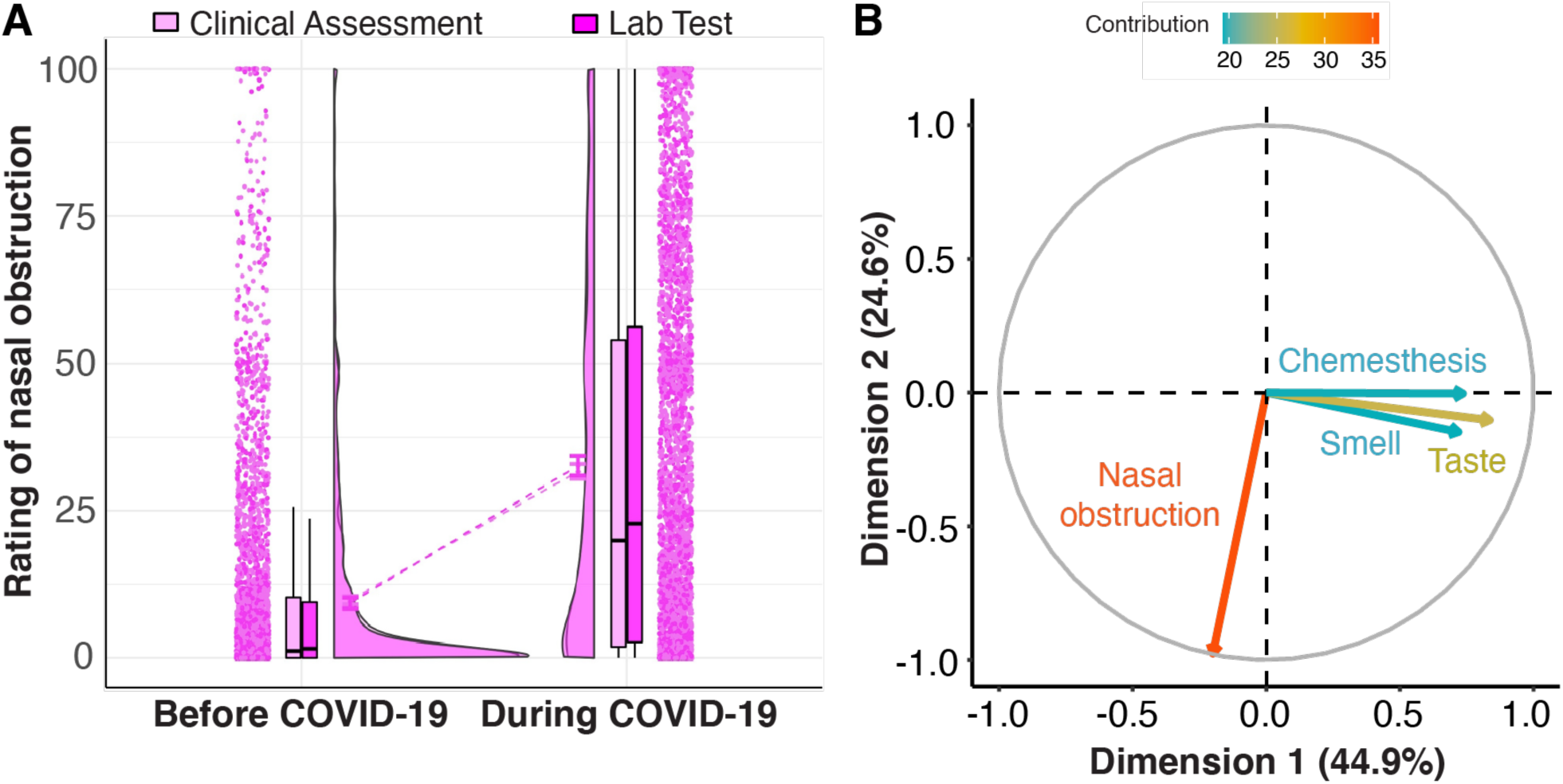
Nasal obstruction. A) The raincloud plot represents ratings for perceived nasal obstruction. From left to right, ratings from single participants are displayed as dots. Boxplots show the 1st to 3rd quartiles, the horizontal line denotes the median, and whiskers denote 1.5 times the interquartile range. The density distribution of the data shows the proportions of given ratings. COVID-19 diagnosis is color-coded, with Clinical Assessment in lighter shade and Lab Test in darker shade. B) Principal component analysis. Correlation circle of the perceptual changes with the 1st (abscissa) and 2nd (ordinate) principal components (PCs).

To further characterize potential relationships between changes in perceived nasal obstruction and reports of changes in the three chemosensory modalities, we computed a Principal Component Analysis (Figure 4B). Changes in smell, taste, and chemesthesis ratings (during minus before) correlated strongly with component 1 (smell: r = 0.72; taste: r = 0.84; chemesthesis: r = 0.74), which explained 45.2% of the total multidimensional variance (inertia). By contrast, change in perceived nasal obstruction was strongly anti-correlated (r = −0.97) with the orthogonal component 2, which explains 24.6% of the total inertia. These results indicate statistical independence of changes in chemosensory ability and perceived nasal obstruction. That is, changes in chemosensory ability and perceived nasal obstruction are statistically independent, so we conclude that changes in olfactory function in COVID-19 positive individuals cannot be attributed to nasal obstruction.

### Chemosensory clustering

Overall, distinct patterns of chemosensory dysfunction/distortion existed among the study participants. We used *k-means* algorithm to cluster respondents based on the similarities and differences in smell, taste, and chemesthesis change (Figure 5). The data-driven, 3-cluster solution (bootstrapped stability = 0.94) identified three groups that can be described by a combination of two chemosensory dimensions: i) the degree of smell and taste loss and ii) the degree of chemesthesis loss. Cluster 1 (N = 1767, blue) is characterized by ratings reflecting substantial smell, taste loss and preserved chemesthesis (centroids: smell: −87.81, taste: −65.97, chemesthesis: −11.07). Cluster 2 (N = 1724, orangr) is characterized by ratings reflecting moderate smell/taste loss and unaffected chemesthesis (centroids: smell: −24.33, taste: −20.97, chemesthesis: −6.87). Cluster 3 (N = 548, green) is characterized by ratings reflecting substantial smell, taste, and chemesthesis loss (centroids: smell: −88.89, taste: −86.74, chemesthesis: −72.39).

**Figure 5.**
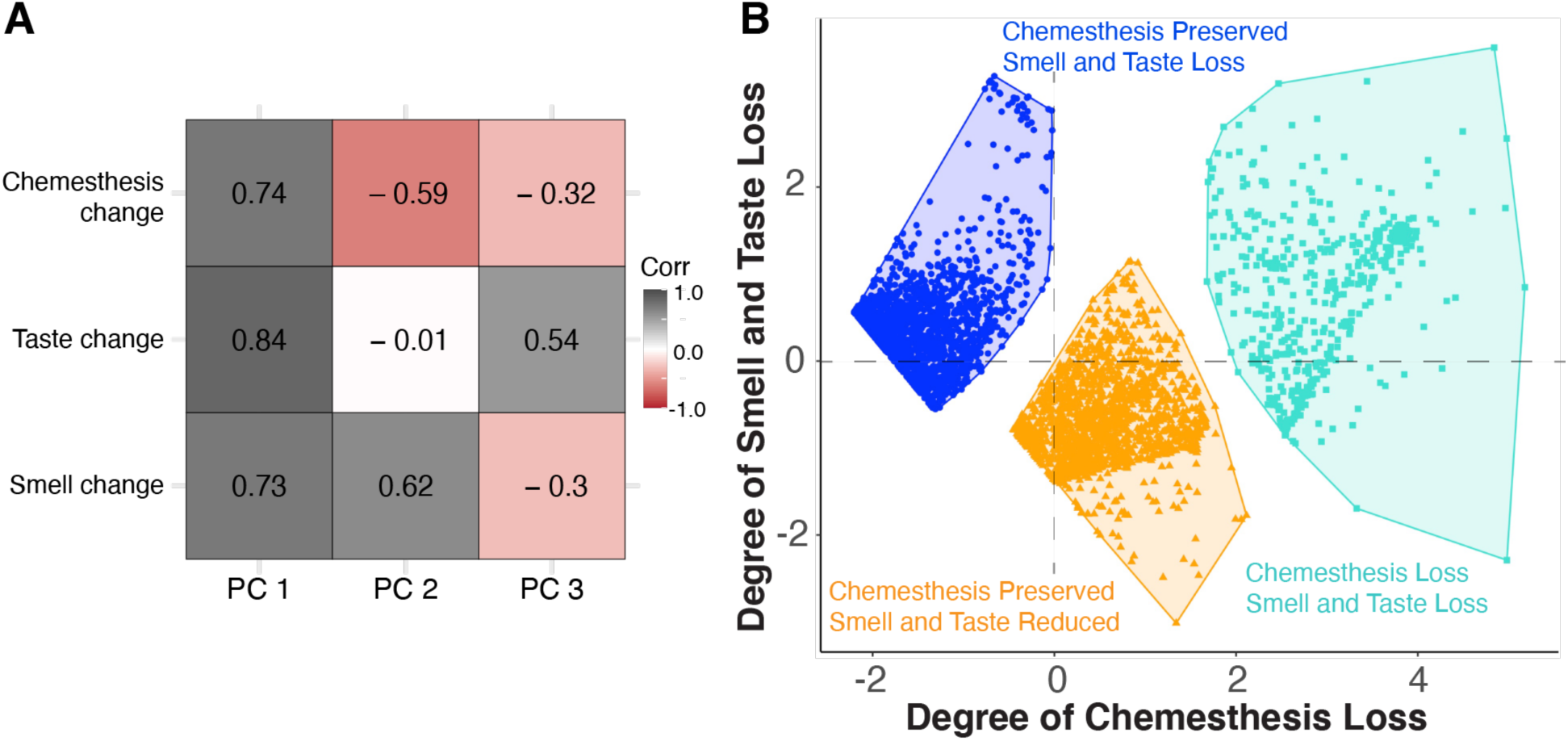
A) Correlations between the three principal components with respect to changes in three chemosensory modalities (i.e. taste, smell, and chemesthesis). Shades of gray indicate positive correlation, whereas shades of red indicate negative correlations. White denotes no correlation. B) Clusters of participants identified by *k-means* clustering. The scatterplot shows each participant’s loading on dimension 1 (degree of chemesthesis loss, abscissa) and dimension 2 (degree of smell and taste loss, ordinate). Loadings for participants in cluster 1 (blue, N=1767) are characterized by significant smell and taste loss and preserved chemesthesis. Participants in cluster 2 (orange, N=1724) are characterized by ratings reflecting moderate smell/taste loss and preserved chemesthesis. Loadings for participants in cluster 3 (green, N=548) are characterized by significant smell, taste and chemesthesis loss.

## Discussion

Our study confirms and substantially extends previous reports showing that smell loss and taste loss are associated with COVID-19. Similar to other recent studies (Bagheri et al., 2020; Chen et al., 2020; Gane et al., 2020; Giacomelli et al., 2020; Haldrup et al., 2020; Hopkins et al., 2020; Lechien et al., 2020a, 2020b; Mao et al.,2020; Menni et al., 2020; Moein et al., 2020; Yan et al., 2020a, 2020b), we find that the majority of our participants with COVID-19 reports a severe reduction in the ability to smell as compared to before the onset of that disease. Notably, this smell loss was not associated with self-reported nasal obstruction, consistent with anecdotal reports. Further, we find that qualitative changes in smell (smell distortions or phantoms) were relatively rare. We found that taste, and to a lesser degree chemesthesis, were also significantly impaired for individuals with COVID-19. Together, these results suggest that COVID-19 broadly impacts chemosensory function across multiple sensory modalities, and that disruption of these may be a possible indicator of COVID-19.

This project is distinct from prior studies on the links between chemosensory dysfunction and COVID-19 in that it leverages a massive crowd-sourced, multinational approach to attack this urgent issue, and does so within a collaborative open science framework. This initial report is based on data in 10 languages from 41 countries; since the first tranche of data on April 18, 2020, 18 additional languages have been added on a rolling basis. The multinational, collaborative nature of the GCCR approach also sets it apart from other recently developed tools. Our hope is that an inclusive globally deployed assessment, coupled with publicly accessible data shared under contemporary open science best practices, will serve as a foundation for future work. It is a limitation of this initial snapshot, however, that participants from different countries are not evenly represented. Cultural biases or country-specific manifestations of COVID-19 could potentially impact these results and will be explored by GCCR in future studies. Though our comprehensive self-report survey cannot replace in-person testing in a controlled clinical or laboratory setting, the gold standard for assessing alterations in chemosensory function, it efficiently and effectively addresses an emerging public health crisis with global scope of coverage. Thus, the model shown in this study of remote smell and taste assessment utilizing the internet may represent one way of reducing delays in assessment until aggressive physical distancing ends (Patel, 2020; Workman et al., 2020).

The mean change in ability to smell was substantial. Prior to onset of COVID-19, the mean rating for the ability to smell was over 90 on a 100-point VAS, yet during the disease, the mean rating dropped below 20. These data do not allow us to differentiate between individuals with partial (hyposmia) versus total loss (anosmia), and participants themselves may be unable to precisely characterize their degree of loss in the absence of objective olfactory testing (Hoffman et al., 2016; Loetsch and Hummel, 2019; Welge-Lüssen et al., 2005). Still, we can conservatively conclude that a major drop in the ability to smell is a hallmark of COVID-19. If the prevalence of COVID-19-associated smell loss is greater than that reported for the common cold or influenza (Beltrán-Corbellini et al., 2020), a different mechanism for disrupting olfactory function may be at play, or this difference could also reflect increased tropism of SARS-CoV-2 for olfactory tissues (Baig et al., 2020).

Critically, the self-reported smell loss we observed is statistically independent of self-reported nasal obstruction. In common URIs, nasal obstruction can explain temporary smell impairments, a phenomenon many individuals have experienced in daily life. Here, estimates of nasal obstruction were based solely on self-report (we asked participants to rate the amount of “nasal blockage”); our data do not include objective, clinically validated measures of nasal breathing or obstruction. While nasal congestion does occur with COVID-19, it appears to be relatively rare in our sample. Still, the fact that many of our participants report substantial loss of olfactory function in the absence of concomitant nasal blockage seems remarkable.

In other instances of post-viral smell loss, about half of patients also experience a qualitative change in smell (Frasnelli et al., 2004; Reden et al., 2007; Rombaux et al., 2009). By contrast, less than 10% of participants in this study reported parosmia or phantosmia symptoms. The rarity of qualitative changes in smell may be a hallmark of COVID-19- associated smell impairments. Alternatively, the present study may not have fully captured qualitative changes in smell, as they tend to emerge later in the course of other disorders (Bonfils et al., 2005) and the present assessment was limited to within at most two weeks of suspected illness or diagnosis. Further studies are needed to more comprehensively address this issue.

While taste loss has also been associated with COVID-19 in patient anecdotes and a few studies, in most cases it has not been clearly differentiated from changes in smell. Here, we found that ratings of taste function were, like those for smell, substantially decreased in individuals with COVID-19. Participant ratings for taste function dropped from a mean of ~ 91 before COVID-19 onset to less than ~24 during the disease. It is well established that people often confuse changes in retronasal olfaction – an important component of flavor perception during eating and drinking – with a true taste loss. While we cannot rule this out completely given the study design, ~60% of those reporting a taste loss also reported a decrease in their perception of at least one specific taste quality, with salty taste being the most common selection. The question on taste qualities is a CATA (check all that apply) question, which means that the subjects can choose any taste qualities that they believe were clearly affected. Indeed, many of the participants chose multiple taste qualities. These data support an interpretation that at least some participants were properly discerning taste from flavor. The observation that some participants reported loss of only a subset of taste qualities may reflect their difficulty in correctly identifying and naming individual taste qualities (Pilkova et al., 1991; Welge-Luessen et al., 2011) rather than quality-selective hypogeusia/ageusia (e.g., Gudziol and Hummel, 2007; Henkin et al., 1970; Lugaz et al., 2002; Huque et al., 2009). However, these possibilities cannot be clarified with the present database.

Compared to smell, the literature has described fewer examples of post-viral taste loss (Adour, 1994; Rubin and Daube, 1999). As the number of people responding to this questionnaire continues to grow on a rolling basis, the differences among different types of respiratory illnesses and their relationship to the degree of taste loss will be a major focus of forthcoming analyses.

Perhaps our most surprising finding was a notable loss of oral chemesthesis ability with COVID-19. Though the decrease is not as large as seen for smell and taste – an ~46% rating reduction for chemesthesis as compared to ~89% and ~76% percentage drop in smell and taste, respectively – it is significant. Interestingly, impairment of chemesthesis was typically accompanied by either taste and smell loss, while taste and smell loss could appear with normal chemesthesis. While nasal chemesthesis experienced with the inhalation of noxious chemicals like ammonia or ethanol is sometimes confused with smell, oral chemesthesis responses to compounds like capsaicin from chili peppers or menthol from mint rarely is (Green, 1996). Though predominantly thought of as the chemical activation of trigeminal afferents carrying temperature, pain or vibration information from the oral, nasal and eye mucosa, other somatosensory nerves, including in the mouth, can also be affected (Green, 1996; McDonald et al., 2016). Chemesthesis (and taste) has been reported to accompany post-viral hyposmia resulting from a URI, at least in some cases (Ren et al., 2012; de Haro-Licer et al., 2013; Pellegrino et al., 2017; Fark and Hummel, 2013). Together with our findings for smell and taste, these data suggest that SARS-CoV-2 impacts all three major chemosensory modalities. The mechanisms are not clear and may be distinct for each chemosensory system. For example, transcriptomic studies of the olfactory mucosa of mouse and human suggests that sustentacular, Bowman’s gland, microvillous cells and stem cell populations, not olfactory sensory neurons themselves, contain ACE2, a receptor required for SARS-CoV-2 viral entry into cells. (Brann et al., 2020; Fodoulian et al., 2020). The pattern of ACE2 expression indicates SARS-CoV-2 may infect tongue keratinocytes (Venkatakrishnan et al., 2020) but it is not known if taste receptor cells or cranial nerves carrying taste or chemesthetic information can be infected by SARS-CoV-2. This virus could alternatively infect surrounding epithelia or blood vessels (Sungnak et al., 2020; Varga et al., 2020), or perhaps even target cells of the central nervous system (Baig et al., 2020).

Based on the stark changes in ratings reported here, one may speculate that both smell and taste loss in COVID-19 are all-or-none phenomena. Although, we cannot rule out that this is an artifact of scale usage, this explanation seems unlikely, as the distribution of the chemesthetic ability ratings is roughly rectangular. This suggests that the all-or-none effect observed for smell cannot be simply attributed to participants using the scale in a discrete rather than continuous fashion. The self-reporting of olfactory function has been used in numerous studies; however, it is not unanimously accepted as it may suffer from low validity (Landis et al., 2003) due to under- and overreporting biases (Dalton and Hummel, 2000; Oleszkiewicz et al., 2020) and possible arbitrary usage. These studies all indicate that self-report ratings are far from being completely inaccurate, especially in participants with severe hyposmia or anosmia, with reported accuracy rates of 70-80% (Lötsch et al., 2019, Hoffman et al., 2016; Rawal et al., 2015).

Here, we account for well-known individual differences in baseline chemosensory abilities, as well as use of rating scales, by using a within-subject design where participants rate their abilities for different time points (before and during COVID-19). We perform an analysis of differences between two assessments (e.g. during minus before COVID-19) rather than on absolute ratings. To better address the question of validity of change in ability ratings, future studies should compare these self-reported and recalled ratings to validated clinical tests before and during the individual’s respiratory illness. However, in times of pandemic, the advantages of a remote assessment method may outweigh the potential decrease in validity compared to face-to-face clinical measures of taste and smell. Still, we acknowledge that a convenience sample recruited online may not be representative of the general population; thus, our study and others that use this type of recruitment approach (e.g., Menni et al., 2020; Iravani et al., 2020) should not be used to estimate prevalence of chemosensory loss in individuals with COVID-19.

Lastly, we found that mean impairments of smell, taste, and or chemesthesis did not differ between study participants who reported a COVID-19 diagnosis based on a Lab Test and those who reported diagnosis based on a Clinical Assessment. However, the Clinical Assessment group exhibited a larger variance in chemosensory loss than the Lab Test group. This could reflect more variability in the accuracy of the diagnosis, as the Clinical Assessment group may include individuals who were misdiagnosed and actually have another viral illness and/or a milder form of the disease. Determining whether the degree of change in chemosensory ability differs between COVID-19-positive individuals and those who are COVID-19-negative but have another respiratory disease will require specific comparisons between those two groups in a future study.

## Conclusions

The GCCR consortium shows how health professionals, clinicians, patient advocates, and scientists can work together to undertake large-scale ground-breaking research of acute public health significance. The present research sets an example of how an emergent response to a global pandemic can be tackled with a crowd-sourced initiative that combines rigorous scientific standards with open-science practices. The established network, research infrastructure, protocol, and findings have the potential to influence current theories on the effects and mechanisms of COVID-19 on the chemical senses and to fuel future research in other areas.

## Data Availability

The data will be available at the OSF project: osf.io/a3vkw

## Acknowledgements

This work was supported financially with discretionary funds from the Pennsylvania State University (Penn State), including a gift from James and Helen Zallie given in support of Sensory Science at Penn State. The authors also wish to thank Paule V. Joseph for her continuous support and for thoughtful discussions as well as Jacqueline Dysart and Karen Phipps at Compusense and Olivia Christman at Penn State for all their help in rapidly deploying the GCCR survey in multiple languages. We are grateful to Marek Vondrak for their help with programming the automatization of the authorship list, Jae-Hee Hong for their editing contribution, and Tristam Wyatt for his role of facilitator of communication among the authors. Additionally, we would like to thank in their role as translators: Aditi Prasad, Alexandros Delides, Ali Khorram-Toosi, Aline Pichon, Amin Homayouni, Amol P Bhondekar, Angela Bassoli, Anshika Singh, Antti Knaapila, Arijit Majumdar, Caterina Dinnella, Debarka Sengupta, Diana Wieck-Fjaeldstad, Dripta Roy, E. Bignon, Eman Hussien Ali Moussa Aboumoussa, Erminio Monteleone, Evangelia Tsakiropoulou, Francesca Boscolonata, Garmt Dijksterhuis, Gaurav Ahuja, Gauri Gharpure, Geetha GT, Giorgia Sollai, Hhardik Shah, Hinal Kharva, Hyoshin Kim, Ingrid Ekström, Ivan Mendez, Jakob Henriksen, Janina Seubert, Jens Sundbøll, Jian Zou, Jitendra Gosai, Kazushige Touhara, Kruttika Phalnikar, Lester Clowney, Lijo Kurian, Marcelo Antonio, Marina Litvak, Mohammad Yaqoob, Musa Ayman Nammari, N. Ravel, Nafiseh Alizadeh, Nasera Rizwana, Neva Bojovic, Nitindra Nath Bandyopadhyay, Orietta Calcinoni, Pavlos Maragoudakis, Pia Soee, Pooja Sarin, Poonam Adhikari, Prasad Kshirsagar, Pratheek HP, Rahul Kottath, Rashid Al Abri, Robert Greene, Rumi Iwasaki, Sanal Aman, Sangyeon Lim, Santosh Rajus, Sara Spinelli, Saurabh Mahajan, Seo Jin Cheong, Shima Taallohi, Simon Singh, Soumya Palit, Sreejith Shankar, Srimanta Pakhira, Sudeshna Bagchi, Sudhir Verma, Takaki Miwa, Takushige Clowney, Tatiana Laktionova, Tatjana Abaffy, Vinaya Sahasrabuddhe, Vinod K Lokku, Xiaojing Cong, Yeonwoo Park, Yiqun Yu, Young Eun, Yuko Nakamura, Zaid Kamal Madni.

